# Social support & sustained physical activity during COVID-19 pandemic

**DOI:** 10.1101/2021.03.04.21252466

**Authors:** Verity Hailey, Abi Fisher, Mark Hamer, Daisy Fancourt

## Abstract

COVID-19 lockdown introduced substantial barriers to physical activity, providing a unique ‘natural experiment’ to understand the social factors associated with sustained physical activity.

**Objectives:** The objectives of this study were to identify the proportion of people who successfully sustained physical activity during lockdown and to explore whether social support, loneliness and social isolation were associated with maintenance of physical activity during COVID-19 lockdown

**Method:** Longitudinal data from the COVID-19 Social Study was used to identify a sample of participants who maintained their physical activity despite lockdown.

**Results:** 16% were consistently active while 44% were completely inactive. After adjustment for multiple confounders high social support was associated with a 39% (95% CI, 12-74%) increased odds of sustaining physical activity. Associations between physical activity and loneliness and social isolation were not found.

**Conclusion:** This study supports previous research showing the importance of social support for the long-term maintenance of physical activity behaviour but shows that such effects extend to contexts of social restrictions.

## Introduction

In response to COVID-19 quarantine strategies such as lockdown, non-essential travel restrictions and social distancing were implemented in an attempt to reduce the spread of the virus (1). The strategies are likely to have impacted the level and patterns of physical activity (PA) (2–4), with potential harmful effects on physical and mental health (1). For example, in the United Kingdom (UK), gyms, leisure facilities and sports clubs were closed, affecting many usual exercise behaviours (5). The pandemic led to major changes in commuter patterns, with many people working from home, furloughed or losing work, reducing active commuting (6). Further, schools and childcare centres were closed, so home-based caring responsibilities increased for many, decreasing the need to walk or travel (5). Results from a systematic review of 66 articles looking at changes in PA during the COVID-19 pandemic showed the impact of such policies, with the majority of studies reporting a decrease in PA (7). Despite restrictions, time outside to exercise was allowed, engaging in daily exercise was encouraged and meeting recommended daily activity levels was possible. Yet not everybody engaged in regular PA. The pandemic restrictions therefore provide a ‘natural experiment’ to explore social determinants of PA behaviour. Identifying factors that are associated with successfully sustaining sufficient levels of activity despite significant barriers could help inform interventions and future pandemic responses.

To date, several studies have focused on *individual predictors* of decreases in PA. A UK smartphone-based tracking study (n=5395) found a larger drop in PA during the lockdown amongst younger people and those who had been active prior to lockdown (8); a finding echoed in a study of 532 Australian students (9). However, other studies have found different results. A cross sectional online study in Belgium (n=13,515) reported that those aged <55 years and were inactive prior to lockdown were likely to exercise more (10). Of note, the mode of usual exercise appeared to be a key factor, with those who usually exercised with friends/sports clubs and who did not engage with online exercise tools reporting a reduction in exercise (10). A study using the COVID-19 Social Study looking at trajectories of PA in relation to lockdown measures found that although 62% experienced little change, nearly 29% reduced PA and 12% of those who didn’t change were consistently inactive (11). A retrospective observational study of 48,440 adults who were diagnosed with COVID-19 showed that people who consistently met the PA guidelines prior to the pandemic were associated with a reduced risk of severe COVID-19 outcomes. Those who were consistently inactive had a greater risk of hospitalisation, admission to intensive care and death (12). To date, the majority of studies exploring predictors of changes in PA have been cross-sectional in nature and used a limited number of variables as predictors.

There is a lack of data to date exploring how individual social factors could have affected changes in PA during the pandemic. *Social support* (defined as the extent to which individuals perceive those around them are available to them and are attentive to their needs) has been associated with a positive effect on well-being (13), PA participation (14) and sustained PA prior to the COVID-19 pandemic (15). Social support is not a single entity but a multi-layered and complex construct (16). There are five main types of social support; instrumental, emotional, informational, companionship and validation support (17), all are associated with PA (16,18).

Stressful events may require multiple resources and types of support (13). There are two main constructs of social support, structural and functional support (16). Structural support describes the existence of relationships and relates to the size, type and frequency of a social network. Functional support relates to the degree to which these relationships serve a function and provide resources; emotional (e.g. companionship and empathy) and instrumental (e.g. financial, practical help) (16). The effect of social support can be explained by two major hypothesis. The stress-buffering hypothesis, where it is thought social support can buffer the negative impact of stressful life events, and the direct-effect hypothesis, where social support has a positive effect on health independent of stress levels (13). People with high social support show overall better health in their daily lives (19). There is also a distinction between actual support received in the past and perceived availability of support. Perceived social support is regarded as a sensitive measure in the context of ability to cope with challenges and is important In predicting its effects on physical and mental health and quality of life (16), perceived social support is used in the paper.

Social support may be particularly important during the pandemic as it has been shown to play a key role in general well-being and is a strong predictor of resilience following disasters and other pandemics (20). Specifically, social support may serve as a ‘buffer’ as per the stress-buffering hypothesis, providing emotional and psychological support, which is considered a major factor in maintaining well-being and coping with health challenges (21). The importance of social support in relation to PA is well understood (14,15,22). The ongoing importance of social support has yet to be explored during the challenges of the pandemic. There is evidence that it might influence other health behaviours. For example, a cross sectional study of changes in alcohol consumption in 1958 US university students (after COVID-19 related campus closure) showed those with greater perceived social support reported less alcohol consumption than those with lower social support (23).

Other social factors, including *social isolation* and *loneliness*, have also been related to PA pre pandemic and levels may have increased as a result of lockdown restrictions. Social isolation and loneliness are distinct from, although related to, social support. While social isolation refers to a lack of social contact with others, loneliness refers to the perception that one’s social contact is insufficient to meet one’s emotional needs (24,25). Social isolation has been shown to have a negative effect on the amount of overall physical activity, with an increase in social isolation directly related to reduced PA (26,27). Loneliness has also been identified as an independent risk factor for a reduction in activity and discontinuation of PA (28).

During the COVID-19 pandemic, social factors such as social isolation, loneliness and social support have all been affected. Quarantine and social distancing have led to elevated levels of loneliness and social isolation (29). Cross sectional results from the UK based COVID-19 Psychological Wellbeing Study showed that rates of loneliness where high with a prevalence of 27% during the initial phase of lockdown (30), with the COVID-19 Social Study reporting a prevalence of 14% for severe loneliness (31). Whether changes in individual-level experiences of social factors such as isolation, loneliness and social support have affected PA remains unknown. Therefore, the aims of this study were to identify the proportion of people who successfully sustained PA during lockdown and to explore whether social support, loneliness and social isolation were associated with maintenance of PA during COVID-19 lockdown. We hypothesized that high social support would be favourably associated with PA, but loneliness and social isolation would have a negative impact on sustained activity.

## Methods

### Study design and participants

Data was used from the COVID-19 Social Study, a large scale longitudinal panel study of adults living in the UK during the COVID-19 pandemic (33). The study is not random and therefore not representative of the UK population but contains a heterogeneous sample (33). Study participation required, a valid email address, internet access, be aged ≥18 years and living it the UK. Recruitment was undertaken using three primary approaches. Firstly, the study was promoted through existing networks (mailing lists, print/digital media coverage and social media. Second, targeted recruitment was undertaken using advertising and recruitment companies focusing on a) low-income backgrounds b) no, or low qualifications c) unemployed. Third, promotion via partnerships with third sector organisations to vulnerable groups. The study commenced on 21st March 2020, and 69,475 people completed at least one week of the 8 weeks included in this study (see supplementary Table 1). Data collection was via a weekly online questionnaire. Baseline data was collected at wave 1 (wave 1 = the week participants join the study), there were questions repeated weekly and one-off modules on a variety of topics. As online data collection, completion of every question was required for submission. The study was approved by UCL Research Ethics Committee (12467/005), with all participants giving informed consent.

Full documentation of data collection protocol is available at www.covidsocialstudy.org/results

## Measures

### Dependent variable

#### Physical activity

Physical activity was self-reported on a weekly basis. Self-report questionnaires are the most common method of PA assessment: they are easy and accurate at measuring intense activity although less robust at measuring light to moderate activity (34). A ‘*stylised questions*’ and ‘*time diaries*’ approach (35) was used to measure ‘time use’ of a specific set of activities including PA (36). Participants were asked to focus on the last weekday, and report how much time they spent on three levels of PA. PA patterns are known to be different between weekends and weekdays, with PA lowest on Sundays and highest on Saturday in some studies (37), however the average amount of time spent in moderate physical activity was not found to be significantly different between week days and weekends (38).

The three categories were; gentle PA (e.g. walking), moderate or vigorous physical activity (MVPA) (e.g. running, cycling, swimming) or exercise inside your home or garden (e.g. yoga, weights, indoor exercise). Time spent doing the different activities was reported as; none, <30 mins, 30mins-2 hours, 3-5 hours and 6+ hours.

Current World Health Organisation (WHO) and UK guidance recommend adults should aim to be active daily and achieve 150minutes of moderate activity per week (39). Benefits of PA are seen at even modest levels of activity such as walking and gardening for 30 min/day on most days of the week. Taking the description of moderate activity into account, the moderate/high intensity and in-home activity categories were combined to identify all those who would have achieved any kind of moderate activity levels. Those who reported <30mins the previous working day were felt unlikely to achieve the recommended 150mins/week and were designated as ‘inactive’, those who reported >30mins were ‘active’. A description of long-term physical activity engagement was generated using a Physical Activity Pattern Index which consists of three ordered categories; inactive, intermittently active, and active. This approach was similar to the one used in a longitudinal study that categorised physical activity levels of middle aged women over 15 years and explored autonomous motivation and self-efficacy with long-term patterns of physical activity behaviour (40). In this study participants were categorized as active or inactive at each of the 8 timepoints. Those who did not report active behaviour at any time point were classified as ‘inactive, those categorised as ‘active’ 1-5 out of the 8 weeks were classified as ‘intermittently active’. Those categorised as ‘active’ 6-8 out of the 8 weeks (≥75%) were classified as ‘active’ as maintenance of physical activity isn’t a continuous behaviour, but a level that is significantly in the intended direction (41)

### Independent variables

#### Social support

Although social support can be measured in a number of different ways, perceived social support is the most commonly measured index (42). In this study perceived social support was measured using the Perceived Social Support Questionnaire (F-SozU K-6) adapted for use in COVID-19 and reported weekly (see supplemental Table 2). This is a 6-item questionnaire with a 5-point Likert scale ranging from 1=not at all, to, 5=very true. The scores for each measure were then summed to give a total ranging from 6 to 30, where the higher the score, indicates higher levels of social support. The questionnaire had excellent validity and reliability for perceived social support (43) with an internal consistency of 0.89 (44) and a Cronbach’s alpha of 0.86 (45) reported in previous studies. Recent research (45) looking at COVID-19 have grouped people into three levels of support, scores of 6-17 = low, 18-25 = normal and 26-30 = high. Categorisation into three levels of support is easier to interpret than a continuous scale of support. This categorisation was adopted in this study.

#### Loneliness

Loneliness was measured using the UCLA-3 loneliness scale, a short form of the Revised UCLA Loneliness Scale (UCLA-R) and reported weekly. It is designed to measure subjective feelings of loneliness as well as feelings of social isolation, it is reliable and has strong validity (46). This is a 3-item scale, respondents were asked how often they felt (1) they lack companionship (2) left out (3) isolated from others. Frequencies ranged from hardly ever (score = 1), some of the time (score = 2) and often (score = 3). The scores of each scale were summed to give a final score ranging from 3 to 9 where higher scores indicated higher levels of loneliness. Researchers in the past have grouped people into categories (26), score of 3-5 = not lonely and scores of 6-9 = lonely.

#### Social isolation

Due to the fast moving nature of the lockdown and survey setup, the social isolation variable was only collected from week 4. Participants were asked what their current isolation status was in line with Government guidelines. Only those who selected that they were in full isolation, not leaving their home, only interacting with their household, were categorised as socially isolated.

### Covariates

We included data on various demographics: gender (male/female), age (18-29, 30-45, 46-59, 60+), ethnicity (white vs BAME [black, Asian and minority Ethnic]), a household income of >£30,000 p/a (yes/no), university education (yes/no), employment (full-time, part-time employment or self-employed vs in education, unable to work, unemployed, homemaker or retired). Data were also collected on living alone (yes/no), urban living (living in a city or town vs living in a village or hamlet), physical health condition such as high blood pressure, diabetes, heart disease (yes/no), having a diagnosed mental health condition including depression, anxiety or any other mental health problem (yes/no), carer status (yes/no), key worker (yes/no), active prior to lockdown (undertaking moderate to vigorous physical activity for ≥15 minutes on 5-7 days in the week prior to lockdown vs on just 0-4 days in the week prior to lockdown).

## Statistical Analysis

Analysis were carried out using Stata 14.0 (StataCorp, College Station, TX). Participants that completed weeks 1 to 8 were included in our analysis.

We examined baseline characteristics of the total population and the two subsamples of active and inactive participants. Chi-squared test was used to identify differences between those that completed the full 1-8 weeks data collection and those that completed only weeks 1-4. Levels of social support, loneliness and social isolation were examined in relation to physical activity status. For the purpose of analysis, PA and covariables were each dichotomized. Physical activity was categorised as inactive and active after review of the Physical Activity Index. The original inactive and intermittent groups were merged into the inactive group due to the high number of participants (23%) in the ‘intermittent’ category who were active for 1-2 weeks and were therefore mostly inactive. Logistic regressions were performed in which PA was regressed individually onto all covariables; demographic (gender, age, ethnicity, income, education level & employment status), health (physical health condition, mental health condition, active prior to lockdown), living condition (lives alone, urban living) and other (carer or key worker). Logistic regression was performed in which PA with loneliness, PA with social support, and PA with social isolation were regressed on 4 models to identify if they influenced PA behavior. Model 1 adjusting for age and gender, model 2 additionally adjusting ethnicity, employment, income and education, model 3 additionally adjusting for physical and mental health conditions, model 4 additionally adjusting for active prior to lockdown.

## Results

### Descriptive analysis

14,485 participants completed the first 4 weeks (weeks 1-4) of which 6906 (48%) participants completed the full 8 weeks and comprised the sample for analysis in the current study. Participant characteristics are presented in Table 1. 74% were female, mean age was 52.7 years (95% CI 52.4 – 53.1) and 96% were white. The sample were well educated with 70% achieving degree level or above education, 60% were employed, and 60% reported a higher income (above £30k) threshold. Key workers accounted for 20% of the participants and 15% were carers. 52% reported a chronic long-term health condition, 42% stating a physical health condition and 17% a mental health condition, 25% reported being active prior to lockdown. Gender, education, and carer responsibility did not have an effect on study response rate. Significant determinants of non-response were found to be, younger age, BAME ethnicity, being a high earner, living in an urban environment, being employed or a key worker, living with others, having any chronic health condition (less likely to complete with mental health condition and more likely to complete having a physical health condition), and being less active prior to lockdown.

**Table 1:** Baseline characteristics of participants (n=6906) who completed data collection weeks 1-8

| Variable | Total Sample<br>(n = 6906) |  | Active<br>(n = 1115) |  | Inactive<br>(n = 5791) |  |
| --- | --- | --- | --- | --- | --- | --- |
|  | n | % | n | % | n | % |
| **Gender** |  |  |  |  |  |  |
| Female | 5116 | 74.1 | 849 | 76.1 | 4267 | 74.7 |
| Male | 1758 | 25.5 | 262 | 23.9 | 1466 | 25.3 |
| **Age category** |  |  |  |  |  |  |
| 18-29 | 438 | 6.3 | 77 | 6.9 | 361 | 6.2 |
| 30-45 | 1762 | 25.5 | 312 | 28 | 1450 | 24 |
| 46-59 | 2206 | 31.9 | 367 | 32.9 | 1839 | 31.8 |
| 60+ | 2500 | 36.2 | 359 | 32.2 | 2141 | 37 |
| **Ethnicity** |  |  |  |  |  |  |
| White | 6652 | 96.3 | 1058 | 94.9 | 5594 | 97 |
| BAME | 254 | 3.7 | 57 | 5.1 | 96.6 | 5.1 |
| **Income** |  |  |  |  |  |  |
| >£30K/year | 3726 | 59.9 | 701 | 70.6 | 3025 | 57.9 |
| <£30K/year | 2493 | 40.1 | 292 | 29.4 | 2201 | 42.1 |
| **Education level** |  |  |  |  |  |  |
| Degree or above | 4804 | 69.6 | 872 | 78.2 | 3932 | 67.9 |
| None/ high school | 2102 | 30.4 | 243 | 21.8 | 1859 | 32.1 |
| **Employed** |  |  |  |  |  |  |
| Yes | 4109 | 59.5 | 723 | 64.8 | 3386 | 58.5 |
| **Key worker** |  |  |  |  |  |  |
| Yes | 1359 | 19.7 | 200 | 17.9 | 1159 | 20 |
| **Carer** |  |  |  |  |  |  |
| Yes | 1065 | 15.4 | 164 | 14.7 | 901 | 15.6 |
| **Lives alone** |  |  |  |  |  |  |
| Yes | 1546 | 22.4 | 219 | 19.6 | 1327 | 22.9 |
| **Urban living** |  |  |  |  |  |  |
| Yes | 5333 | 77.2 | 894 | 80.2 | 4439 | 76.7 |
| **Chronic health condition** |  |  |  |  |  |  |
| Yes | 3591 | 52 | 410 | 36.8 | 2905 | 50.2 |
| **Physical health condition** |  |  |  |  |  |  |
| Yes | 2924 | 42.3 | 344 | 30.9 | 2580 | 44.6 |
| **Mental health condition** |  |  |  |  |  |  |
| Yes | 1152 | 16.7 | 129 | 11.6 | 1023 | 17.7 |
| **Active prior to lockdown** |  |  |  |  |  |  |
| Yes | 1746 | 25.3 | 531 | 47.6 | 1215 | 21 |

The physical activity index, see Table 2, shows that 44% of participants were inactive, 40% were intermittently active and 16% were consistently active. Within the intermittently active group, the majority (59%) were active for only 1 or 2 weeks. Fewest of the intermittent group (12%) were active for 5 weeks. Within the active group, there was a fairly even split of those active for 6, 7 or 8 weeks. Table 2 provides full details of physical activity index and within category results.

**Table 2.** Number of active weeks and Physical Activity Index

| Number of active weeks | Number of participants | Total group % | Within category % |
| --- | --- | --- | --- |
| **Inactive N=3043 (44.1%)** |  |  |  |
| 0 | 3043 | 44.1% | 100% |
| **Intermittently active N=2748 (39.8%)** |  |  |  |
| 1 | 964 | 14% | 36% |
| 2 | 629 | 9.1% | 23% |
| 3 | 398 | 5.8% | 14% |
| 4 | 436 | 6.3% | 15% |
| 5 | 321 | 4.6% | 12% |
| **Consistently active N=1115 (16.1%)** |  |  |  |
| 6 | 348 | 5% | 31% |
| 7 | 375 | 5.4% | 34% |
| 8 | 392 | 5.7% | 35% |
| **Total** | **6906** | **100** |  |

Association with persistent PA behaviour, with no adjustment for covariates, was found with social support, being BAME, having higher income, being employed, having a university level education. Factors that were adversely associated with PA included loneliness, social isolation, living alone, having a physical or mental health condition, being active prior to lockdown and being aged 60+ (Table 3).

**Table 3.** Effects of covariables on sustained physical activity

| **Physical Activity** |  |
| --- | --- |
| **Variable** | **Odds ratio (95% CI)** |
| **Social support** |  |
| Low | Reference |
| medium | 1.31 (1.08 – 1.59)** |
| high | 1.74 (1.44 – 2.11)*** |
| **Loneliness** |  |
| low | Reference |
| high | 0.82 (0.71 – 0.95)** |
| **Social isolation (>wk 4)** |  |
| Yes | 0.72 (0.02 – 0.55)* |
| **Gender** |  |
| Male | Reference |
| female | 1.14 (0.98 – 1.32) |
| **Age** |  |
| 18-29 | 0.99 (0.75 – 1.3) |
| 30-45 | Reference |
| 46-59 | 0.93 (0.79 – 1.09) |
| 60+ | 0.77 (0.19 – 0.92)** |
| **Ethnicity** |  |
| White | Reference |
| BAME | 1.53 (1.13 – 2.07)** |
| **High income** |  |
| Yes | 1.75 (1.51 – 2.02)*** |
| **Employed** |  |
| Yes | 1.31 (1.14 – 1.5)*** |
| **University education** |  |
| Yes | 1.7 (1.46 – 1.98)*** |
| **Lives alone** |  |
| Yes | 0.82 (0.7 – 0.96)* |
| **Urban environment** |  |
| Yes | 1.23 (1.05 – 1.44)* |
| **Physical health condition** |  |
| Yes | 0.56 (0.48 – 0.64)*** |
| **Mental health condition** |  |
| Yes | 0.61 (0.5 – 0.74)*** |
| **Carer** |  |
| Yes | 0.94 (0.78 – 1.12) |
| **Key worker** |  |
| Yes | 0.87 (0.74 – 1.03) |
| **Active prior to lockdown** |  |
| Yes | 3.42 (3-3.91)*** |

\*P<0.05; \*\*P<0.01; \*\*\*P<0.001

### Social support

There was an association between PA and social support. Mean social support score was ‘normal’ and ranged from 22.59 (SE 0.07) in week 1, with a slight decrease to 22 (SE 0.09) in week 8. Of those who were active, 14% had low social support, 41% had medium support and 45% had high support. Those who were inactive, 20% had low support, 44% had medium and 36% had high support. Logistic regression demonstrated an increase in likelihood of being active amongst individuals with both medium and high support (Table 3). High social support continued to be positively associated with PA, OR 1.39 (95% CI 1.12 – 1.74) p=0.003 even when accounting for all demographic, health-related factors and other covariates (Table 4). Medium social support was only associated when accounting for age and gender, OR 1.71 (95% CI 1.41 – 2.08) p=0.009 and then lost with the addition of further covariates (Table 4).

**Table 4.** Effects of social support, loneliness and isolation on sustained physical activity. N=6906.

| **Physical Activity** |  |
| --- | --- |
| **Variable** | **Odds ratio (95% CI)** |
| **Social support** |  |
| Model 1 |  |
| medium | 1.29 (1.07 – 1.57)** |
| high | 1.71 (1.41 – 2.08)*** |
| Model 2 |  |
| medium | 1.18 (0.96 – 1.46) |
| high | 1.54 (1.24 – 1.9)*** |
| Model 3 |  |
| medium | 1.12 (0.91 – 1.38) |
| high | 1.42 (1.15 – 1.76)** |
| Model 4 |  |
| medium | 1.14 (0.92 – 1.42) |
| high | 1.39 (1.12 – 1.74)** |
| **Loneliness** |  |
| Model 1 |  |
| high | 0.79 (0.69 – 0.92)** |
| Model 2 |  |
| high | 0.86 (0.74 – 1.00) |
| Model 3 |  |
| high | 0.93 (0.8 – 1.09) |
| Model 4 |  |
| high | 0.95 (0.81- 1.12) |
| **Social isolation** |  |
| Model 1 |  |
| Yes | 0.76 (0.58 – 0.99)* |
| Model 2 |  |
| Yes | 0.84 (0.62 – 1.13) |
| Model 3 |  |
| Yes | 1.01 (0.74 – 1.37) |
| Model 4 |  |
| Yes | 1.24 (0.91 – 1.69) |

\***P<0.05**; \*\***P<0.01**; \*\*\***P<0.001** Model 1: adjusted for age and gender. Model 2: additionally adjusted for ethnicity, employment status, income and education. Model 3: additionally adjusted for chronic physical and mental health conditions. Model 4: additionally adjusted for active prior to lockdown

### Loneliness

Mean loneliness score was ‘not lonely’ and ranged from 4.67 (SE 0.021) in week 1, with a slight reduction in loneliness (showed by an increase in mean score) to 4.8 (SE 0.023) in week 8. Percentage of people reported as lonely, per the UCLA scale, was 31.5% in week 1, increasing to 35.5% in week 8, chi squared test showed no statistical difference in loneliness.

There was a reduction in likelihood of being active amongst people who were lonely, OR 0.82 (95% CI 0.71 – 0.95) p=0.007. Loneliness remained associated with PA after accounting for age and gender, OR 0.79 (95% CI 0.69 – 0.92) p=0.002, although the association was attenuated after further covariate adjustments (Table 4).

### Social isolation

In unadjusted models there was an association between isolation and lower odds of regular PA, OR 0.72 (95% CI 0.02 – 0.55) p=0.017. This association was retained after accounting for age and gender, 0.76 (95% CI 0.58 – 0.99) p=0.041, although attenuated in further models (Table 4).

## Discussion

Management of COVID-19 has created barriers for how people interact and maintain PA. In this large UK-wide study of adults we identified a sub-sample of participants that were able to maintain their PA despite restrictions. Those with high social support had a 39% increased odds of sustaining PA during lockdown. However, although associations between loneliness and social isolation and decreased odds of sustaining PA during lockdown were observed in minimally-adjusted models, this association was lost after adjusting for wider covariates.

When looking cross-sectionally at the data, levels of self-reported physical activity in our study are similar to those from other UK sources. For example, Sport England (2020) reported that 32% of adults were meeting the guidelines of 150mins/week MVPA in the last week of April 2020, whilst our study reported 26% active for the same week. Such levels are concerning as they are lower than the estimated 63-66% of adults who met aerobic guidelines prior to COVID-19 (47). However, our study built on previously reported cross-sectional data by showing that just 16% of adults analysed maintained recommended levels of physical activity throughout lockdown, 44% were inactive and a further 23% were active for only 1 or 2 weeks. This demonstrates the difference in those meeting the guidance when looking cross-sectionally compared to longitudinally and suggests that the number of people who were consistently active during the first UK lockdown was substantially lower than the cross-sectional reports. It is well known that not achieving the recommended levels of aerobic exercise is associated with poor physical health, poor mental health and premature mortality (1). This finding alone suggests that more work needs to be done on supporting people during COVID-19 and potential future pandemics to meet PA guidelines on a regular basis in order to get maximum benefit from the activity.

Our study also explored what predicted the likelihood of an individual engaging in sustained PA across lockdown. Being white, well educated, a high earner, urban living and good health status are all well-known predictors of PA (48,49). Any form of health condition, physical or mental health and older age, are associated with a lower likelihood of being active (49). Our findings were broadly in line with these pre COVID-19 predictors. However, although female gender would normally have a negative impact on PA (49), this was not been seen in our data, with more women (17%) remaining active then men (15%). This results reflects increased PA levels seen amongst women, older adults and those with a long term condition (47).

We focused specifically on social predictors of sustained PA. Social support was found to be a consistent predictor but loneliness and isolation were only associated in less-adjusted statistical models. The reasons for this may have been both direct and indirect. Directly, theories that are commonly used in PA interventions e.g. Social Cognitive Theory, Theory of Planned Behaviour, Socio Ecological Model, and Health Belief all contain social support as a key factor in affecting behaviours (15). The findings reported here suggest that even during social restrictions when such support may be disrupted from usual patterns (e.g. offered virtually rather than face-to-face), social support remains a key influencer of PA behaviours. Indirectly, it is also possible that social support may have played a role in buffering against the negative effects of poor mental health on PA during the pandemic. There is a large literature showing how mental health was adversely affected during the first UK lockdown (50). Poor mental health is associated with lower PA engagement (51). But research during the pandemic suggested that social interactions helped to reduce the experience of depressive symptoms, supporting the findings presented here(52). While the pandemic may have led to rises in loneliness and social isolation, this was situational due to lockdown. Chronic or prolonged social isolation and loneliness has a known negative impact on health and wellbeing (30), but it is possible that short-term loneliness and isolation do not have the same effect. Should there be multiple lockdowns there is potential for the increased rates of loneliness to become chronic leading to it having an impact on physical activity.

The strengths of the study include its longitudinal design. It allows for multiple data points and for us to identify those participants who maintained their PA throughout lockdown. The sample provided information on a range of demographic factors, health conditions and social factors in addition to physical activity behaviours which has given us a unique opportunity to look at social isolation along with social support and loneliness. Limitations of the study include non-random sampling approach leading to a less representative sample of the UK population. As with many studies, participants were highly educated, white and female. The study used self-reported measure of PA leaving it open to reporting bias e.g. imprecise recall. Attempts were made to minimise this by providing examples of common types of exercise with corresponding intensities. Asking participants to self-report on a single day of activity has its limitations, PA was one of thirteen measures of time use/activities which were collected. Due to concerns about focusing on a ‘typical’ day, which involves aggregating information from multiple days and averaging, a ‘time diary’ approach was used based on the previous weekday. There is potential that an ‘active’ person was allocated as ‘inactive’ if they had not undertaken physical activity on the previous working day. However to achieve the WHO guidelines of 150min MVPA/week, regular adherence should be ≥5 days/week of ≥30mins MVPA. It would be unlikely that a participant would be incorrectly coded for all 8 weeks. An ‘active’ participant were those coded as active 75% of the time, allowing for low activity weeks e.g. illness, work/family pressure, lack of activity on the previous weekday. This study looks at those who have remained active throughout lockdown, we are not aware of similar data published anywhere else looking at sustained activity.

The potential for multiple lockdowns over extended periods of time could cause prolonged periods of low PA for a substantial proportion of the population leading to increased risk of issues with physical and mental health. Previous research shows the importance of social support for initiating PA, this study demonstrates the importance of social support for the long-term maintenance of PA behaviour within the context of social restrictions and suggests that it does not need to be delivered face to face. Other social factors, such as loneliness and social isolation, were less consistent with their impact on PA. The development of interventions and programs to support PA both during and outside of pandemic situations should ensure that social support is built in using theories that have shown to promote regular PA participation. The pandemic has prompted the development of virtual and remote PA through online classes and communities, supporting these programs to build in social support could be beneficial to supporting regular PA both now and in the future.

## Data Availability

Anonymous data will be made available following the end of the pandemic.

## Funding

VH was funded by the ESRC-BBSRC Soc-B Centre for Doctoral Training, ES/P000347/1. The Covid-19 Social Study was funded by Nuffield Foundation [WEL/FR-000022583], but the views expressed are those of the authors and not necessarily the Foundation. The study was also supported by the MARCH Mental Health Network funded by the Cross-Disciplinary Mental Health Network Plus initiative supported by UK Research and Innovation [ES/S002588/1], and by the Wellcome Trust [221400/Z/20/Z]. DF was funded by the Wellcome Trust [205407/Z/16/Z]. The researchers are grateful for the support of a number of organisations with their recruitment efforts including: the UKRI Mental Health Networks, Find Out Now, UCL BioResource, SEO Works, FieldworkHub, and Optimal Workshop. The study was also supported by HealthWise Wales, the Health and Care Research Wales initiative, which is led by Cardiff University in collaboration with SAIL, Swansea University. The funders had no final role in the study design; in the collection, analysis and interpretation of data; in the writing of the report; or in the decision to submit the paper for publication. All researchers listed as authors are independent from the funders and all final decisions about the research were taken by the investigators and were unrestricted.

The research questions in the UCL COVID-19 Social Study built on patient and public involvement as part of the UKRI MARCH Mental Health Research Network, which focuses on social, cultural and community engagement and mental health. This highlighted priority research questions and measures for this study. Patients and the public were additionally involved in the recruitment of participants to the study and are actively involved in plans for the dissemination of findings from the study.

## Authors contributions

VH analysed and interpreted the data and was primarily responsible for the drafting of the manuscript. AF & MH contributed to the topic, statistical analysis, interpretation of the findings and revised the manuscript. DF launched the study and contributed to its original concept and design. All authors reviewed, critically appraised and approved the final manuscript.

## References

1. Füzéki E, Groneberg DA, Banzer W. Physical activity during COVID-19 induced lockdown: Recommendations. J Occup Med Toxicol. 2020;15(1):1–5.

2. Ammar A, Brach M, Trabelsi K, Chtourou H, Boukhris O, Masmoudi L, et al. Effects of COVID-19 Home Confinement on Eating Behaviour and Physical Activity : Results of the. Nutrients. 2020;12(1583):13.

3. Chtourou H, Trabelsi K, H’Mida C, Boukhris O, Glenn JM, Brach M, et al. Staying physically active during the quarantine and self-isolation period for controlling and mitigating the covid-19 pandemic: A systematic overview of the literature. Front Psychol. 2020;11(August).

4. Constant A, Conserve DF, Gallopel-Morvan K, Raude J. Socio-Cognitive Factors Associated With Lifestyle Changes in Response to the COVID-19 Epidemic in the General Population: Results From a Cross-Sectional Study in France. Front Psychol. 2020;11(September):1–9.

5. UK Government. COVID-19 restrictions, UK Government [Internet]. 2020 [cited 2020 Feb 3]. Available from: https://www.gov.uk/government/speeches/pm-address-to-the-nation-on-coronavirus-23-march-2020

6. Office for National Statistics. Coronavirus and travel to work : June 2020. 2020;(June):1–22.

7. Stockwell S, Trott M, Tully M, Shin J, Barnett Y, Butler L, et al. Changes in physical activity and sedentary behaviours from before to during the COVID-19 pandemic lockdown : a systematic review. 2021;1–8.

8. McCarthy H, Potts HWW, Fisher A. Physical Activity Behavior Before, During, and After COVID-19 Restrictions: Longitudinal Smartphone-Tracking Study of Adults in the United Kingdom. J Med Internet Res [Internet]. 2021 Feb 3;23(2):e23701. Available from: https://www.jmir.org/2021/2/e23701

9. Gallo LA, Gallo TF, Young SL, Moritz KM, Akison LK. The impact of isolation measures due to COVID-19 on energy intake and physical activity levels in Australian university students. medRxiv. 2020;

10. Constandt B, Thibaut E, De Bosscher V, Scheerder J, Ricour M, Willem A. Exercising in times of lockdown: An analysis of the impact of COVID-19 on levels and patterns of exercise among adults in Belgium. Int J Environ Res Public Health. 2020;17(11):1–10.

11. Bu F, Bone JK, Mitchell JJ, Steptoe A, Fancourt D. Longitudinal changes in physical activity during and after the first national lockdown due to the COVID-19 pandemic in England. Sci Rep. 2021 Dec 1;11(1).

12. Sallis R, Young DR, Tartof SY, Sallis JF, Sall J, Li Q, et al. Physical inactivity is associated with a higher risk for severe COVID-19 outcomes: A study in 48 440 adult patients. Br J Sports Med. 2021 Oct 1;55(19):1099–105.

13. Cohen S, Wills TA. Stress, Social Support, and the Buffering Hypothesis. Vol. 98, Psychologkal Bulletin. 1985.

14. Scarapicchia TMF, Amireault S, Faulkner G, Sabiston CM. Social support and physical activity participation among healthy adults: A systematic review of prospective studies. Int Rev Sport Exerc Psychol. 2017;10(1):50–83.

15. Lindsay Smith G, Banting L, Eime R, O’Sullivan G, van Uffelen JGZ. The association between social support and physical activity in older adults: A systematic review. Int J Behav Nutr Phys Act. 2017;14(1):1–21.

16. Kocalevent RD, Berg L, Beutel ME, Hinz A, Zenger M, Härter M, et al. Social support in the general population: Standardization of the Oslo social support scale (OSSS-3). BMC Psychol. 2018;6(1):4–11.

17. Cutrona C, Suhr J. Controllability of Stressful Events and Satisfaction with Spouse Support Behaviors. Communic Res. 1992;19(2):154–74.

18. Stapleton JN, Lox CL, Gapin JI, Pettibone JC, Karen L. Social Support As a Stage Specific Correlate of Physical Activity. Exerc Phys Educ Res [Internet]. 2015;3:63–79. Available from: https://pdfs.semanticscholar.org/1509/a35aca3153d88a4220f3e0dff9b936cd546a.pdf

19. Buchwald P. Social support. Curated Ref Collect Neurosci Biobehav Psychol. 2016;(September 2016):435–41.

20. Saltzman LY, Hansel TC, Bordnick PS. Loneliness, Isolation, and Social Support Factors in Post-COVID-19 Mental Health. Psychol Trauma Theory, Res Pract Policy. 2020;12:55–7.

21. Zysberg L, Zisberg A. Days of worry: Emotional intelligence and social support mediate worry in the COVID-19 pandemic. J Health Psychol. 2020;

22. Molloy GJ, Dixon D, Hamer M, Sniehotta FF. Social support and regular physical activity: Does planning mediate this link? Br J Health Psychol. 2010;15(4):859–70.

23. Lechner W V., Laurene KR, Patel S, Anderson M, Grega C, Kenne DR. Changes in alcohol use as a function of psychological distress and social support following COVID-19 related University closings. Addict Behav [Internet]. 2020;110(June):106527. Available from: https://doi.org/10.1016/j.addbeh.2020.106527

24. Smith KJ, Gavey S, RIddell NE, Kontari P, Victor C. The association between loneliness, social isolation and inflammation: A systematic review and meta-analysis. Neurosci Biobehav Rev [Internet]. 2020;112(September 2019):519–41. Available from: https://doi.org/10.1016/j.neubiorev.2020.02.002

25. de Jong Gierveld J, van Tilburg T. The De Jong Gierveld short scales for emotional and social loneliness: Tested on data from 7 countries in the UN generations and gender surveys. Eur J Ageing. 2010;7(2):121–30.

26. Steptoe A, Shankar A, Demakakos P, Wardle J. Social isolation, loneliness, and all-cause mortality in older men and women. Proc Natl Acad Sci U S A. 2013;110(15):5797–801.

27. Schrempft S, Jackowska M, Hamer M, Steptoe A. Associations between social isolation, loneliness, and objective physical activity in older men and women. BMC Public Health. 2019;19(1):1–10.

28. Hawkley LC, Thisted RA, Cacioppo JT. Loneliness Predicts Reduced Physical Activity: Cross-Sectional & Longitudinal Analyses. Heal Psychol. 2009;28(3):354–63.

29. Hwang TJ, Rabheru K, Peisah C, Reichman W, Ikeda M. Loneliness and social isolation during the COVID-19 pandemic. Int Psychogeriatrics. 2020;32(10):1217–20.

30. Groarke JM, Berry E, Graham-Wisener L, McKenna-Plumley PE, McGlinchey E, Armour C. Loneliness in the UK during the COVID-19 pandemic: Cross-sectional results from the COVID-19 Psychological Wellbeing Study. Murakami M, editor. PLoS One [Internet]. 2020 Sep 24 [cited 2021 Feb 19];15(9):e0239698. Available from: https://dx.plos.org/10.1371/journal.pone.0239698

31. Bu F, Steptoe A, Fancourt D. Loneliness during lockdown: Trajectories and predictors during the COVID-19 pandemic in 35,712 adults in the UK. medRxiv. 2020;

32. Tison Geoffrey H. Annals of Internal Medicine Worldwide Effect of COVID-19 on Physical Activity : Ann Intern Med Intern. 2020;(March):1–3.

33. Fancourt D, Steptoe A, Bu F. Trajectories of depression and anxiety during enforced isolation due to COVID-19: longitudinal analyses of 59,318 adults in the UK with and without diagnosed mental illness. medRxiv [Internet]. 2020;2020.06.03.20120923. Available from: https://doi.org/10.1101/2020.06.03.20120923

34. Sylvia LG, Bernstein EE, Hubbard JL, Keating L, Anderson EJ. Practical guide to measuring physical activity. J Acad Nutr Diet. 2014;114(2):199–208.

35. Seymour G, Malapit HJ, Quisumbing A. Measuring Time Use in Development Settings. Meas Time Use Dev Settings. 2017;(July).

36. Bu F, Steptoe A, Mak HW, Fancourt D. Time use and mental health in UK adults during an 11-week COVID-19 lockdown: a panel analysis. Br J Psychiatry. 2021;1–6.

37. Racette SB, Weiss EP, Schechtman KB, Steger-May K, Villareal DT, Obert KA, et al. Influence of weekend lifestyle patterns on body weight. Obesity. 2008;16(8):1826–30.

38. Buchowski MS, Acra S, Majchrzak KM, Sun M, Chen KY. Patterns of physical activity in free-living adults in the Southern United States. Eur J Clin Nutr. 2004;58(5):828–37.

39. NHS. NHS Physical activity Guidelines [Internet]. NHS. 2021. Available from: https://www.nhs.uk/live-well/exercise/

40. Janssen I, Dugan SA, Karavolos K, Lynch EB, Powell LH. Correlates of 15-year maintenance of physical activity in middle-aged women. Int J Behav Med. 2014;21(3):511–8.

41. Kwasnicka D, Dombrowski SU, White M, Sniehotta F. Theoretical explanations for maintenance of behaviour change: a systematic review of behaviour theories. Health Psychol Rev. 2016;10(3):277–96.

42. Li F, Luo S, Mu W, Li Y, Ye L, Zheng X, et al. Effects of sources of social support and resilience on the mental health of different age groups during the COVID-19 pandemic. BMC Psychiatry. 2021;21(1):1–14.

43. Kliem S, Mößle T, Rehbein F, Hellmann DF, Zenger M, Brähler E. A brief form of the Perceived Social Support Questionnaire (F-SozU) was developed, validated, and standardized. J Clin Epidemiol. 2015;68(5):551–62.

44. Lin M, Hirschfeld G, Margraf J. Psychological Assessment Brief Form of the Perceived Social Support Questionnaire (F-SozU K-6): Validation, Norms, and Cross-Cultural Measurement Invariance in the. Am Psychol Assoc. 2018;

45. Labrague LJ, De los Santos JAA. COVID-19 anxiety among front-line nurses: Predictive role of organisational support, personal resilience and social support. J Nurs Manag. 2020;28(7):1653–61.

46. Tull MT, Edmonds KA, Scamaldo KM, Richmond JR, Rose JP, Gratz KL. Psychological Outcomes Associated with Stay-at-Home Orders and the Perceived Impact of COVID-19 on Daily Life. Psychiatry Res [Internet]. 2020;289(April):113098. Available from: https://doi.org/10.1016/j.psychres.2020.113098

47. Sport England. SURVEY INTO ADULT PHYSICAL ACTIVITY ATTITUDES AND BEHAVIOUR [Internet]. 2020. Available from: https://www.sportengland.org/news/new-exercise-habits-forming-during-coronavirus-crisis

48. Bauman AE, Reis RS, Sallis JF, Wells JC, Loos RJF, Martin BW, et al. Correlates of physical activity: Why are some people physically active and others not? Lancet [Internet]. 2012;380(9838):258–71. Available from: http://dx.doi.org/10.1016/S0140-6736(12)60735-1

49. Smith L, Gardner B, Fisher A, Hamer M. Patterns and correlates of physical activity behaviour over 10 years in older adults: Prospective analyses from the English Longitudinal Study of Ageing. BMJ Open. 2015;5(4):1–5.

50. Fancourt D, Steptoe A, Bu F. Trajectories of anxiety and depressive symptoms during enforced isolation due to COVID-19 in England: a longitudinal observational study. The Lancet Psychiatry. 2021;8(2):141–9.

51. Shor R, Shalev A. Barriers to involvement in physical activities of persons with mental illness. Health Promot Int. 2016;31(1):116–23.

52. Sommerlad A, Marston L, Huntley J, Livingston G, Lewis G, Steptoe A, et al. Social relationships and depression during the COVID-19 lockdown: Longitudinal analysis of the COVID-19 social study. Psychol Med. 2021;

